# Associations of antidepressants and antipsychotics with lipid parameters: Do *CYP2D6*/*CYP2C19* genes play a role? A UK population-based study

**DOI:** 10.1101/2022.05.03.22273718

**Authors:** Alvin Richards-Belle, Isabelle Austin-Zimmerman, Baihan Wang, Eirini Zartaloudi, Marius Cotic, Caitlin Gracie, Noushin Saadullah Khani, Yanisa Wannasuphoprasit, Marta Wronska, Yogita Dawda, David P.J. Osborn, Elvira Bramon

**Affiliations:** Mental Health Neuroscience Research Department, Division of Psychiatry, University College London, London, W1T 7BN, UK; Social, Genetic and Developmental Psychiatry Centre, Institute of Psychiatry, Psychology and Neuroscience, King’s College London, London SE5 8AF, UK; Department of Pharmacy, Central and North West London NHS Foundation Trust, London, NW1 3AX, UK; Epidemiology and Applied Clinical Research Department, Division of Psychiatry, University College London, London W1T 7BN, UK; Camden and Islington NHS Foundation Trust, 4 St Pancras Way, London, NW1 0PE, UK

## Abstract

**Background:** Dyslipidaemia is an important risk factor for cardiovascular morbidity in people with severe mental illness and which contributes to premature mortality in this population. The link between antipsychotics and dyslipidaemia is well-established, whilst evidence on antidepressants is mixed.

**Aims:** To investigate (1) if antidepressant/antipsychotic use was associated with lipid parameters in UK Biobank participants, and (2) if *CYP2D6* and *CYP2C19* genetic variation plays a role.

**Methods:** Review of self-reported prescription medications identified participants taking antidepressants/antipsychotics. Total, low-, and high-density lipoprotein (L/HDL-C) cholesterol and triglycerides derived from blood samples. *CYP2D6* and *CYP2C19* metabolic phenotypes were assigned from genetic data. Linear regression investigated study aims.

**Results:** Of 469,739 participants, 36,043 took antidepressants and 3,255 antipsychotics. Significant associations were found between use of amitriptyline, fluoxetine, citalopram/escitalopram, sertraline, paroxetine, and venlafaxine with worse levels of each lipid parameter (i.e., higher total cholesterol, LDL-C, and triglycerides and lower HDL-C). Venlafaxine was associated with the worst lipid profile (total cholesterol, mean difference: 0·21 mmol/L, 95% confidence interval [CI]: 0·17 to 0·26, *p*<0·001). Antipsychotic use was associated with lower HDL-C and higher triglycerides (0·31 mmol/L, 95% CI 0·28 to 0·35, *p*<0·001). In participants taking sertraline, the *CYP2C19* intermediate metaboliser phenotype was associated with higher HDL-C (0·05 mmol/L 95% CI: 0·01 to 0·09, *p*=0·007) and lower triglycerides (-0·17 mmol/L 95% CI: -0·29 to -0·05, *p*=0·007).

**Conclusions:** Antidepressants are significantly associated with adverse lipid profiles, potentially warranting baseline and regular monitoring of lipids. Further research should investigate why the *CYP2C19* intermediate metaboliser phenotype may be protective for HDL-C and triglycerides in people taking sertraline.

## INTRODUCTION

Dyslipidaemia is an important risk factor for cardiovascular morbidity^1^ in people with severe mental illness^2^ and which contributes to premature mortality in this population.^3^ The link between antipsychotic use and dyslipidaemia is well-established,^4–6^ and United Kingdom (UK) National Institute for Health and Care Excellence (NICE) guidelines recommend monitoring blood lipid profiles (i.e., total, low-, and high-density lipoprotein (L/HDL-C) cholesterol, triglycerides and total cholesterol to HDL-C ratio) in those prescribed them,^7^ whilst evidence and guidance regarding antidepressants are mixed, with a paucity of high-quality studies.^8,9,10,11^

Several relatively small observational studies report associations between antidepressant use and dyslipidaemia,^8,9,10,11^ including higher triglycerides and lower HDL-C,^8,9^ though, in one study, these associations became non-statistically significant when adjusted for additional potential confounders (a significant association with hypercholesterolaemia remained).^9^ Some studies implicate tricyclics as most detrimental,^8^ and others, selective serotonin reuptake inhibitors.^10^ One study linked serotonin high-affinity antidepressants to higher LDL-C and non-high affinity antidepressants to higher triglycerides.^11^ These studies, however, all had limited power to explore the relative effects of individual medications. A 2006 review concluded that certain antidepressants, such as tricyclics and mirtazapine, may negatively impact lipids more so than others (i.e. bupropion, venlafaxine, duloxetine), but noted low methodological quality of included studies and called for robust studies.^12^ The electronic medicines compendium, which provides access to manufacturers’ summaries of product characteristics for UK-licensed medicines, does not list lipid-related reactions for several antidepressants (e.g. amitriptyline, citalopram/escitalopram, fluoxetine), but lists increased cholesterol as common for paroxetine and venlafaxine and rare for sertraline.^13^

Multiple meta-analyses report links between individual antipsychotics and dyslipidaemia.^4–6^ A 2010 head-to-head meta-analysis of second-generation antipsychotics (48 blinded randomised trials), reported that olanzapine led to significantly greater increases in total cholesterol than aripiprazole, risperidone, and ziprasidone and that quetiapine led to greater increases than risperidone.^4^ A 2020 meta-analysis (21 studies), found olanzapine significantly increased triglycerides, total cholesterol and LDL-C, with no significant change in HDL-C.^5^ An overview of systematic reviews found aripiprazole increased cholesterol, but to a lesser extent than olanzapine, with results based on low or moderate quality evidence.^6^ The electronic medicines compendium lists: increased cholesterol and triglycerides as very common for quetiapine and olanzapine (the former also linked to adverse L/HDL-C); increased cholesterol as uncommon, and increased triglycerides rare, for risperidone; hypercholesterolemia and hypertriglyceridemia very rare for clozapine - whilst lipids are not mentioned for prochlorperazine, and lipid changes noted not clinically important for aripiprazole.^13^

Wide inter-individual variation in efficacy and adverse reactions of antidepressants and antipsychotics exists. Pharmacogenetics could play a significant role in individualising drug therapy.^14,15^ The Cytochrome P450 (CYP450) superfamily of enzymes are heavily involved in the metabolism of many prescribed medications;^16^ with CYP2D6 and CYP2C19 heavily involved for antidepressants and antipsychotics. CYP450 enzymes are also essential in the body’s synthesis of cholesterol.^16^ The genes encoding these enzymes are highly polymorphic and thus represent promising pharmacogenetic targets.^14,15^ Individuals can be phenotyped, respectively, based on *CYP2D6* and *CYP2C19* polymorphisms: ‘normal metabolisers’ carry two homozygous wild-type alleles and have normal enzymatic capacity; ‘poor metabolisers’ carry two loss-of-function alleles and have no enzymatic capacity; ‘intermediate metabolisers’ have reduced enzymatic capacity compared to normal metabolisers but greater capacity than poor metabolisers (e.g. one wild-type and one reduced capacity allele); and ‘rapid’ and ‘ultra-rapid metabolisers’ have greater than normal enzymatic capacity, due to either at least one increased function allele or duplications of functional allele(s).^14,15^ Phenotypes, of which distributions vary across ancestries, impact medication plasma concentrations and risk of adverse reactions,^17,18^ with poor metabolisers predicted as most at risk due to greater concentrations.

Few studies have investigated variation in *CYP2D6* and *CYP2C19* and lipid parameters in the context of antidepressant/antipsychotic medications. One study genotyped 150 inpatients with depression (most receiving antidepressants and over half, antipsychotics) for *CYP2D6, CYP2C9* and *CYP2C19* and calculated four combinatory-gene indices, all of which significantly correlated with total cholesterol, LDL-C and HDL-C, but not triglycerides.^19^ A study of 76 patients taking risperidone found a significant negative change in HDL-C from pre-treatment to 8 weeks post-treatment in carriers of *CYP2D6*\*2 and *CYP2D6*\*65.^20^ These studies, however, did not account for use of other relevant medications such as statins, a mainstay of dyslipidaemia treatment (raising the possibility of interactions), and were too small to draw conclusions.

### Aims

Our aims were to investigate (1) if antidepressant/antipsychotic use was associated with lipid parameters in a large sample of participants from UK Biobank, and (2) if *CYP2D6* and *CYP2C19* genetic variation plays a role influencing lipid parameters in participants taking antidepressants/antipsychotics.

## METHODS

### Study design

This population-based, observational, cohort study used genetic and cross-sectional data from UK Biobank^21,22^ - a major biomedical database with around 500,000 participants. UK Biobank received ethical approval from the North West – Haydock Research Ethics Committee (reference: 21/NW/0157). All participants provided written informed consent.

### Participants

UK Biobank methods have been described elsewhere.^21^ We used data from the baseline visit, where, in brief, participants, aged 37-73, attended one of 22 UK assessment centres between 2006-2010 and completed an extensive set of measures, including questionnaires and interviews (e.g. demographics, medical history, medication use), and provided biological samples.

### Outcomes

Lipid parameters investigated were total cholesterol, LDL-C, HDL-C, and triglycerides, measured in millimoles per litre (mmol/L), extracted directly from UK Biobank (originally derived from non-fasting venous blood samples analysed using a Beckman Coulter AU5800). To aid interpretation, we calculated and used the total cholesterol to HDL-C (TC:HDL) ratio (commonly used and clinically informative)^7^ as an additional outcome in analyses addressing aim one.

### Exposures

Exposures were (a) antidepressants/antipsychotics and (b) *CYP2D6* and *CYPC19* genetic metabolic phenotypes. We reviewed self-reported prescription medications data to identify all antidepressants/antipsychotics, considering both generic and proprietary names (identified through multiple sources^23–25^), and combined equivalent medications under the generic name for analyses. We combined citalopram and escitalopram (the active enantiomer of citalopram)^23^ as one medicine for analyses. We investigated aims in individual medications only if reported as being taken by ≥1,800 participants (consistent with a previous study).^18^ Medications not reaching this threshold were considered for inclusion in a higher-level combined group (e.g. all antipsychotics together).

For genetic exposures, we leveraged genome-wide genotyping and processing conducted centrally by UK Biobank.^22^ Genotyping was performed using the Affymetrix UK BiLEVE Axiom array on an initial sample (50,000 participants) and the Affymetrix UK Biobank Axiom® array (Affymetrix, Santa Clara, CA, USA) for all subsequent participants. These arrays include >820,000 variants (with good coverage of pharmacogenetics variants), with subsequent imputation of >90 million variants. Using the fully-imputed dataset, we performed local quality control and assigned CYP450 metabolic phenotypes, as described previously.^18^ In brief, to include and account for participants of non-European ancestry (European ancestry was determined centrally), two rounds of principal component analysis was conducted (using PC-AiR^26^ and PC-Relate^27^), identifying four ancestry groups (East Asian, South Asian, African, admixed with predominantly European origin); participants not clustering with any main group were excluded. Subsequent processing excluded: variants with minor allele frequency <1% and/or Fisher information score of <0.3 in each ancestry group; one of each pair of participants with a kinship score >0·083 (approximately third-degree relatives); and participants with >10% missingness, excessive genetic relatedness (>10 third-degree relatives); or a mismatch between self-reported and genetically-inferred sex.

To assign CYP metabolic phenotypes, we extracted *CYP2D6* and *CYP2C19* regions of interest (defined as one mega-base upstream of the 5′ end and one mega-base downstream of the 3′ end of the gene). Using an input map and reference panel from the 1000 genomes project,^28^ haplotypes were constructed based on genetic data, imputed using Beagle (version 5·0),^29^ according to the star allele nomenclature system.^30^ Haplotypes containing no star allele-defining single-nucleotide polymorphism variants were classified as wild-type (*1) alleles for the corresponding gene. We grouped individuals into *CYP2C19* metabolic phenotypes based on activity of the individual haplotypes and resulting diplotypes^30^ and into *CYP2D6* phenotypes according to the activity score method.^31^ We did not have data on *CYP2D6* copy number variants and were unable to define *CYP2D6* ultra-rapid metabolisers, or other whole gene deletions (e.g., *CYP2D6*\*5).

### Statistical analysis

Aim one analyses considered all participants with data on at least one lipid parameter and, from these, aim two, participants with high-quality genetic data.

In addition to lipid parameters, across each medication group - we described participants’ age at recruitment (years), sex, self-reported ethnic background, body mass index (BMI) (kg/m2), selected self-reported illnesses (depression, anxiety, schizophrenia, bipolar disorder) and concomitant use of cholesterol lowering medications (ascertained from participants’ response of “cholesterol lowering medications” to the multiple-choice question “do you regularly take any of the following medications?”). We reported proportions for “healthy” versus “not healthy” categories of each lipid parameter (using UK National Health Service categories).^32^ We included a comparison group of participants not taking antidepressants/antipsychotics. We presented lipid parameters stratified by concomitant cholesterol lowering medication status. In participants with high-quality genetic data, we reported distributions of genetically-determined ancestry group, *CYP2D6* and *CYP2C19* metabolic phenotypes and use of strong/moderate inhibitors (as per United States Food and Drug Administration).^33^ Means with standard deviations (SD), medians with interquartile ranges (IQR) and/or counts and proportions were used, as appropriate.

We ran two linear regression models for each lipid parameter as a continuous outcome. The first, using all participants, investigated associations of use of each medication (main predictor) with each lipid parameter (outcome), adjusted for age (continuous), sex (binary) and use of cholesterol lowering medication (binary). The second investigated the pharmacogenetic associations of *CYP2D6* and/or *CYP2C19* metabolic phenotype (main predictor(s)) with each lipid parameter (outcome). These models included participants with high-quality genetic data and were run in each medication group. *CYP2D6* and *CYP2C19* were modelled together where both genes are majorly involved in the metabolism of the medication (as per Clinical Pharmacogenetics Implementation Consortium),^14,15,34^ with normal metaboliser phenotypes used as reference. In addition to age, sex, and cholesterol lowering medication, these models were adjusted for genetic ancestry group (categorical) and, as relevant, use of strong/moderate *CYP2D6* or *CYP2C19* inhibitors (binary). We adjusted for strong/moderate inhibitors (e.g., paroxetine and quinidine for CYP2D6 and fluoxetine and fluvoxamine for CYP2C19), as they play a major role in phenoconversion, a phenomenon whereby drug-gene or drug-drug-gene interactions may result in an observed phenotype different from the genetically-predicted phenotype.^35^ We did not adjust for BMI in order to avoid the risk of overadjustment bias.^36^

Effect estimates are reported with 95% confidence intervals (CIs) and uncorrected *p* values in tables and forest plots. Given analysis of three independent (LDL-C, HDL-C and triglycerides), and two highly correlated/combinatory (total cholesterol and TC:HDL ratio (the latter not included in pharmacogenetic analyses)), outcomes, we corrected for multiple testing by using an adjusted significance threshold of <0·013 (i.e., 0·05/4). Analyses were conducted in Stata/MP version 17·0 (StataCorp LLC).

## RESULTS

### Participants

Overall, 469,739 participants had data for at least one lipid parameter. Of these, 36,043 reported taking at least one antidepressant, 3,255 at least one antipsychotic, whilst 431,853 did not take either. Use of both an antidepressant and an antipsychotic was reported by 1,412 participants. A wide range of medications were reported - with amitriptyline, citalopram, and fluoxetine the most common antidepressants and prochlorperazine, olanzapine and quetiapine the most common antipsychotics (Figure 1). Amitriptyline, fluoxetine, citalopram/escitalopram, paroxetine, sertraline, and venlafaxine met sample size threshold for individual medication analyses. No individual antipsychotic met threshold; all reported antipsychotics were therefore analysed together. Of participants taking antidepressants/antipsychotics, 33,472 had high-quality genetic data.

**Figure 1.**
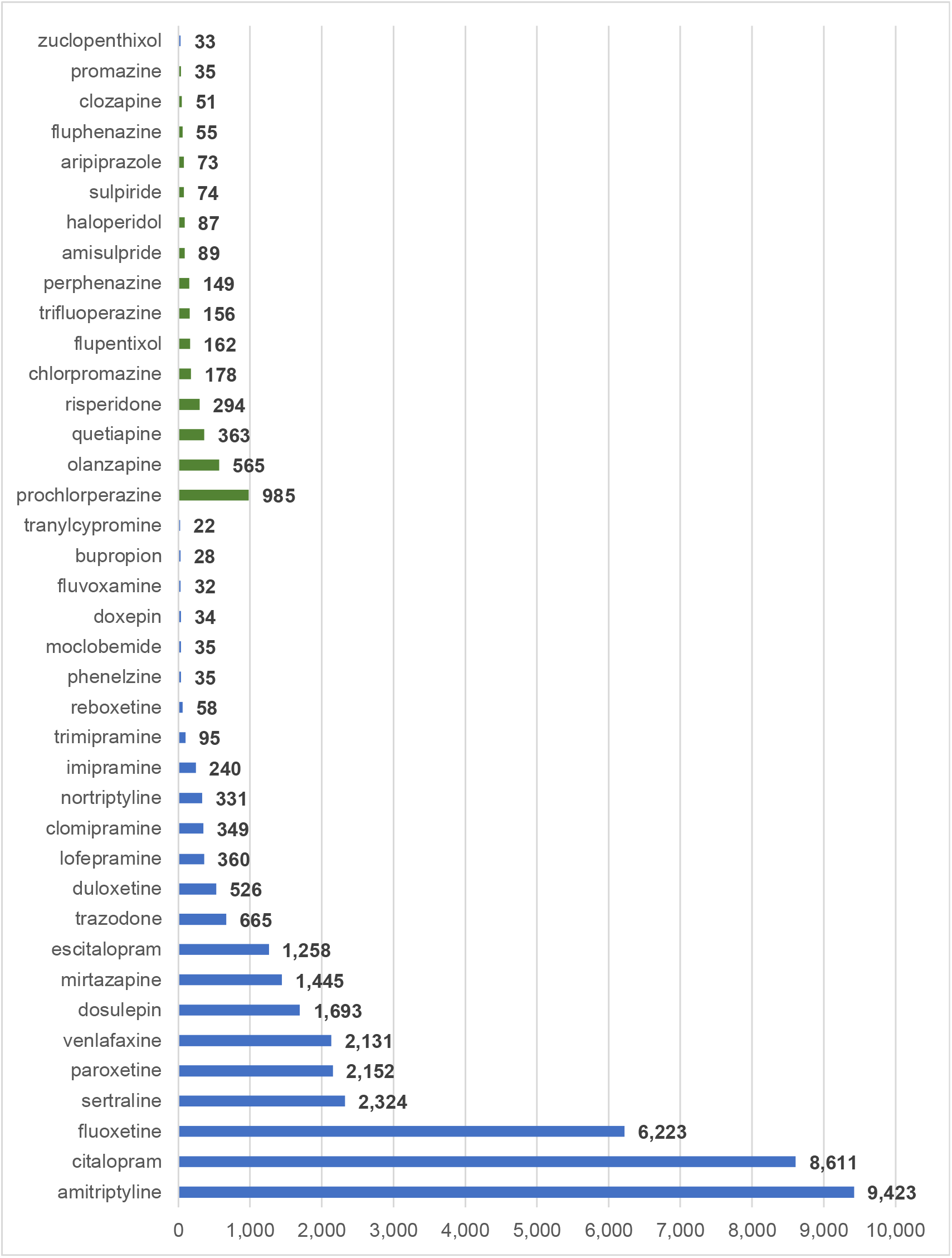
Frequency of antipsychotic and antidepressant drug use in UK Biobank participants. Numbers refer to the number of participants taking each medication. Antidepressants are shown in purple and antipsychotics in green. Medications were self-reported and are included if reported by at least 20 participants.

Sample characteristics, including demographics, unadjusted lipid parameters and genetic metabolic phenotypes, are shown in Table 1 and Supplementary Table 1. Compared to participants not taking antidepressants/antipsychotics, median age was similar across medication groups, but the proportion of females was consistently higher. Median BMI was highest in participants taking venlafaxine (28·4 kg/m2) and antipsychotics (28·2 kg/m2). Across antidepressants, participants taking amitriptyline had the lowest proportion of self-reported depression (17.5%) and anxiety (4.6%), whilst 27.9% taking antipsychotics self-reported schizophrenia or bipolar disorder. Nearly a quarter (24·7%) of participants taking antidepressants/antipsychotics were also taking cholesterol lowering medications, compared to around a sixth (16·8%) in those not taking antidepressants/antipsychotics. Unadjusted lipid parameters stratified by cholesterol lowering medication status are shown in Supplementary Table 2. For *CYP2D6*, most (26,154, 71.4%) participants were normal metabolisers, with 1,919 (5.2%) and 8,585 (23.4%) poor and intermediate metabolisers, respectively. For *CYP2C19*, most were either normal (13,939, 38.0%) or intermediate (10,860, 29.6%) metabolisers, with 1,255 (3.4%) poor and 10,604 (28.9%) rapid or ultra-rapid metabolisers.

**Table 1.**
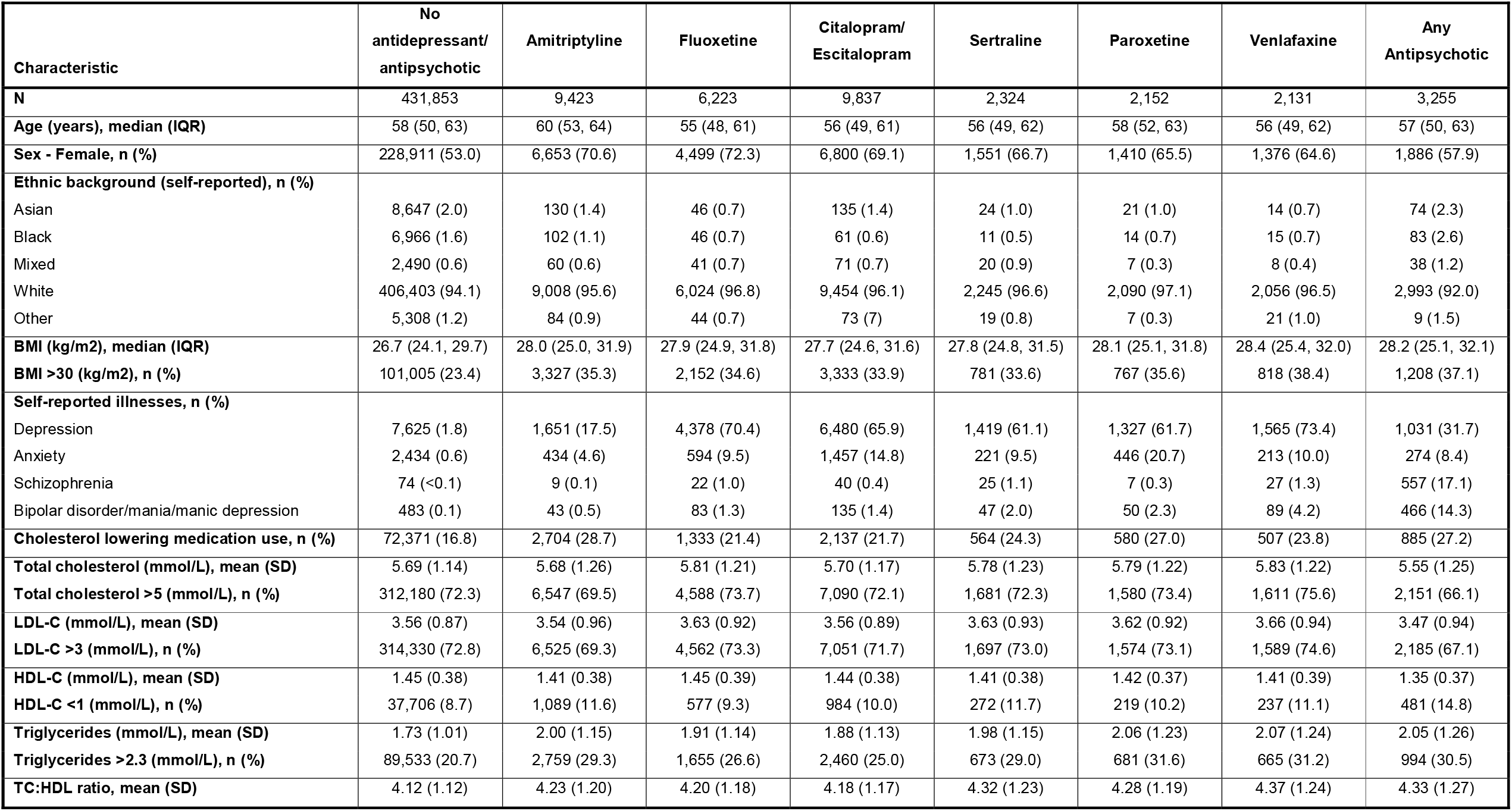
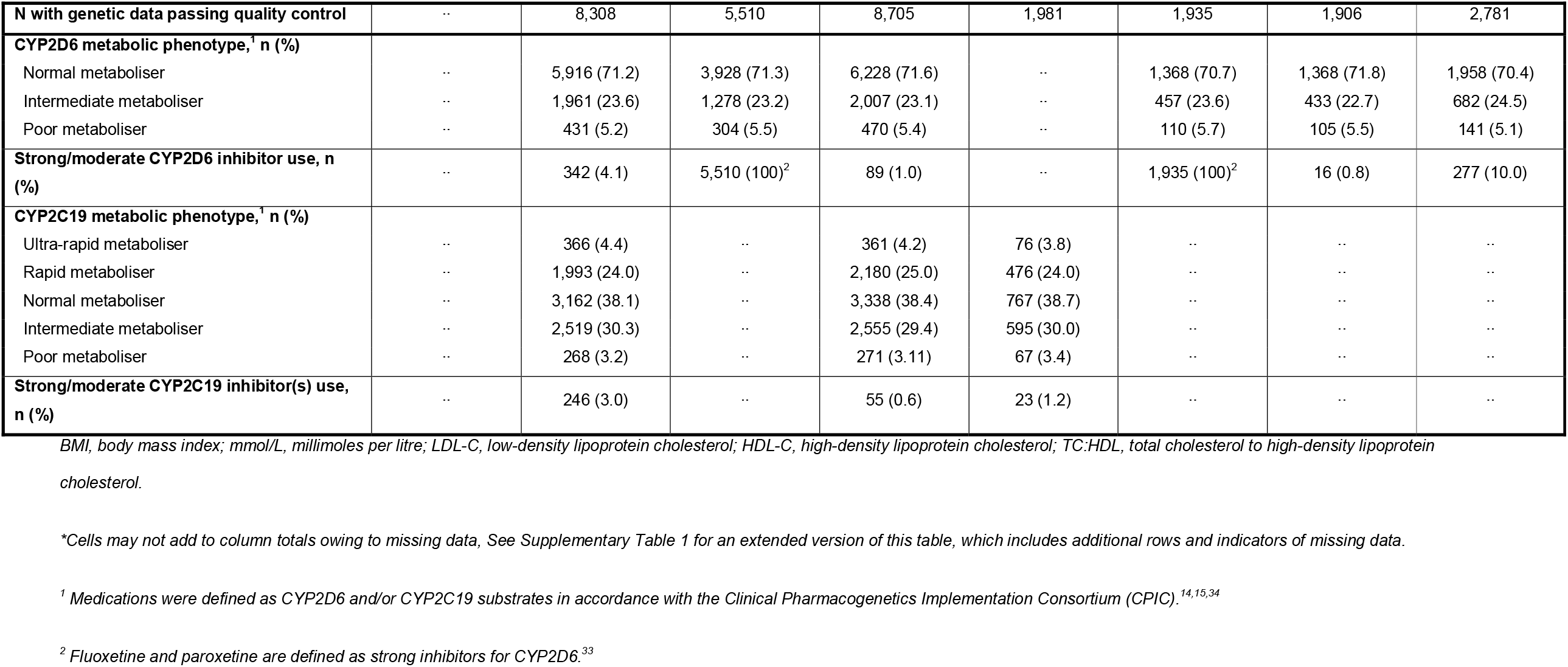
Sample characteristics.*

### Antidepressants, antipsychotics and lipid parameters

Significant associations were found with the use of each antidepressant and each lipid parameter, respectively, when compared to participants not taking the medication (Figure 2, Supplementary Table 3). Antipsychotic use was significantly associated with lower HDL-C (mean difference: -0·10 mmol/L, 95% CI: -0·11 to -0·08; *p*<0.001) and higher triglyceride levels (0·31 mmol/L, 95% CI 0·28 to 0·35; *p*<0·001), but not with total cholesterol or LDL-C.

**Figure 2.**
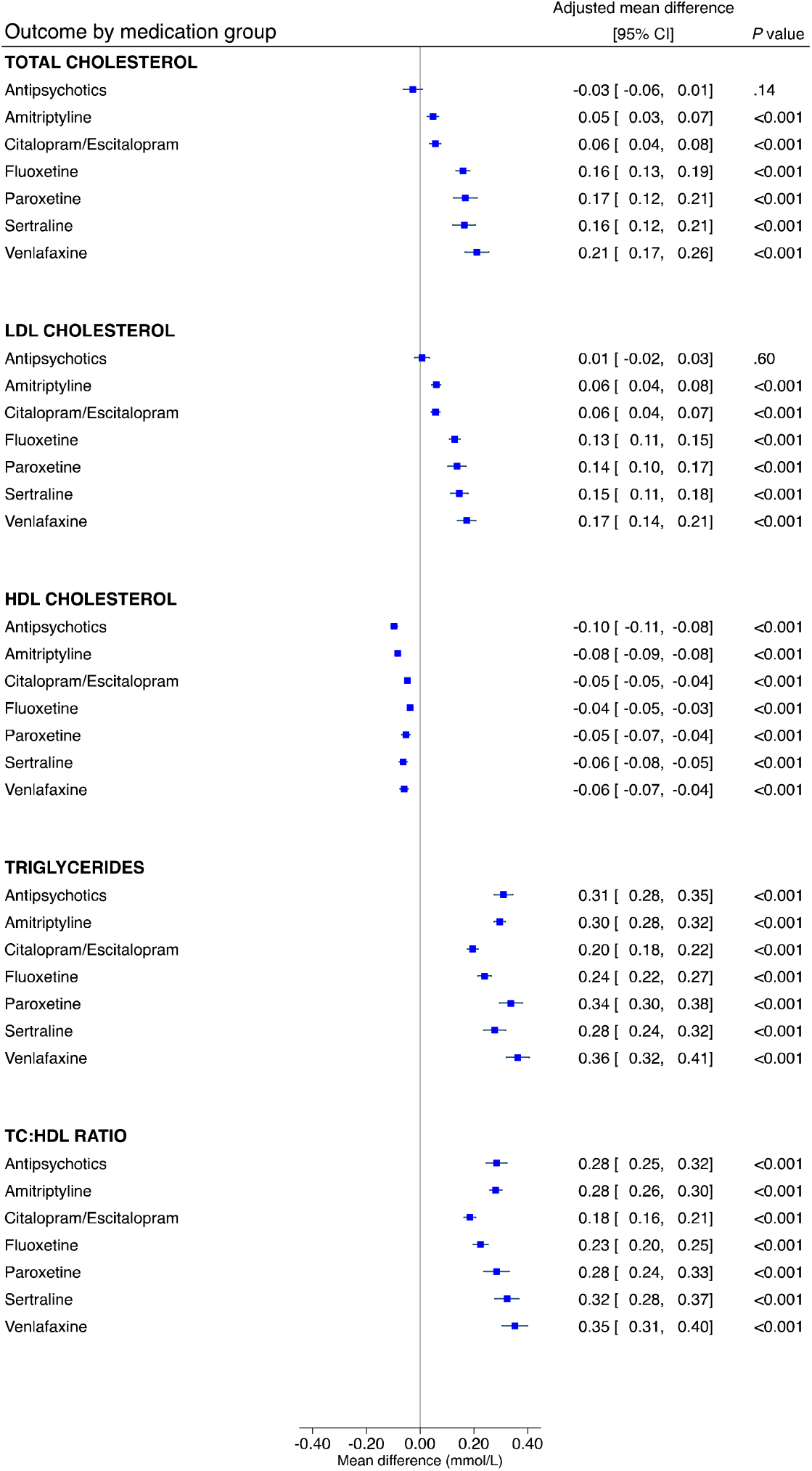
The associations between antidepressants and antipsychotics and lipid parameters. LDL, low-density lipoprotein; HDL, high-density lipoprotein; TC:HDL, total cholesterol to high-density lipoprotein cholesterol ratio; mmol/L, millimoles per litre. Linear regression models were adjusted for age, sex and concomitant use of cholesterol lowering medications; effect estimates are coefficients for the main predictor variable, which was a binary variable defined by whether participants were taking the relevant drug (or not). A total of 469,591 participants contributed total cholesterol data, 468,708 for LDL cholesterol, 429,873 for HDL cholesterol and 469,216 for triglycerides.

Venlafaxine was associated with the highest levels of total cholesterol (mean difference: 0·21 mmol/L, 95% CI: 0·17 to 0·26; *p*<·001), followed by paroxetine (0·17 mmol/L, 95% CI: 0·12 to 0·21; *p*<·001) and sertraline (0·16 mmol/L, 95% CI: 0·12 to 0·21; *p*<·001). A similar pattern was observed for LDL-C. The lowest HDL-C levels were observed with antipsychotics and with amitriptyline (-0·08 mmol/L, 95% CI: -0·09 to -0·08; *p*<·001). The highest triglyceride levels were observed with venlafaxine (0·35 mmol/L, 95% CI: 0·31 to 0·40; *p*<·001) and sertraline (0·32 mmol/L, 95% CI: 0·28 to 0·37; *p*<·001).

To explore the impact of the most frequently reported antipsychotic (prochlorperazine) in the grouped “all antipsychotics” analyses, we conducted a post-hoc analysis excluding prochlorperazine and results were consistent with the primary analyses (data not shown).

### The influence of CYP2D6 and CYP2C19 metabolic phenotypes

Adjusted estimates of the influence of *CYPD6* and *CYP2C19* metabolic phenotypes on lipid parameters across each medication group are shown in Supplementary Table 4 and Table 2, respectively. In participants taking sertraline, the *CYP2C19* intermediate metaboliser phenotype was significantly associated with an average 0·05 mmol/L higher HDL-C (95% CI: 0·01 to 0·09, *p*=0·007) and with an 0·17 mmol/L lower triglyceride level (95% CI: -0·29 to -0·05, *p*=0·007), compared with normal metabolisers (Figure 3). As significant associations were in an unanticipated direction, we undertook post-hoc analyses exploring the impact of cholesterol lowering medications (Supplementary Table 5). Extension of the sertraline HDL-C model to include an interaction term for *CYP2C19* metabolic phenotype by cholesterol lowering medications resulted in a main effect of the intermediate metaboliser phenotype larger than the primary analysis (0·08 mmol/L, 95% CI: 0·03 to 0·12, *p*=0·001), with an intermediate metaboliser phenotype by cholesterol lowering medications interaction effect in the opposite direction (-0·10 mmol/L, 95% CI: -0·19 to -0·01, *p*=0·03, n=126). Stratified analysis revealed a statistically strong effect of the *CYP2C19* intermediate metaboliser phenotype in participants not taking cholesterol lowering medications (0.08 mmol/L, 95% CI: 0.03 to 0.12, *p*=0·001, n=424), but no evidence in those taking them. There was no evidence of an interaction in the extended triglycerides model, but some evidence in stratified analysis of a stronger association in participants not taking cholesterol lowering medications (-0.15 mmol/L, 95% CI: -0.29, -0.02, *p*=0·03, n=456).

**Table 2.**
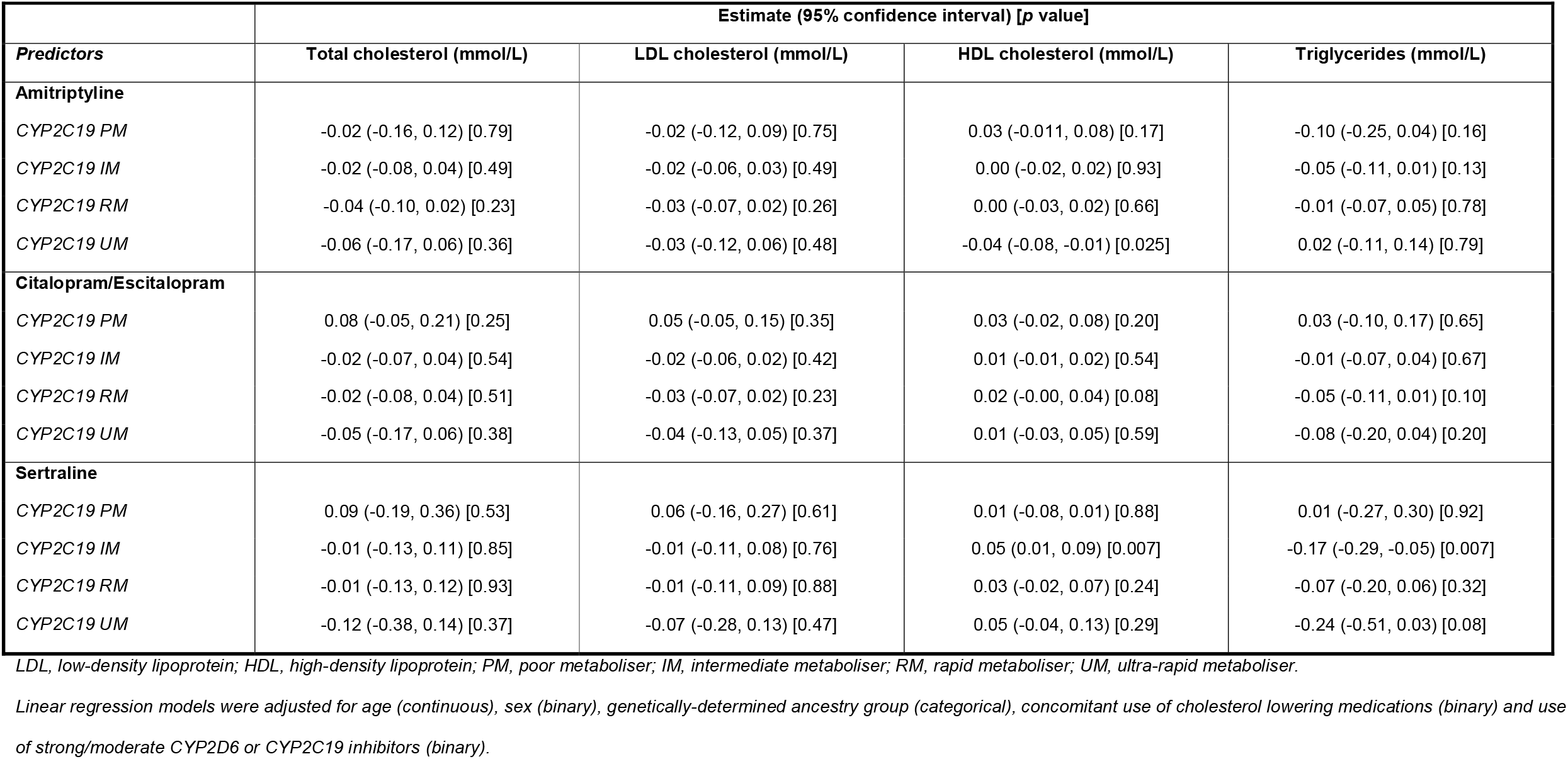
The influence of CYP2C19 metabolic phenotypes on lipid parameters in participants taking antidepressants or antipsychotics.

|  | Estimate (95% confidence interval) [p value] |  |  |  |
| --- | --- | --- | --- | --- |
| Predictors | Total cholesterol (mmol/L) | LDL cholesterol (mmol/L) | HDL cholesterol (mmol/L) | Triglycerides (mmol/L) |
| **Amitriptyline** |  |  |  |  |
| CYP2C19 PM | -0.02 (-0.16, 0.12) [0.79] | -0.02 (-0.12, 0.09) [0.75] | 0.03 (-0.011, 0.08) [0.17] | -0.10 (-0.25, 0.04) [0.16] |
| CYP2C19 IM | -0.02 (-0.08, 0.04) [0.49] | -0.02 (-0.06, 0.03) [0.49] | 0.00 (-0.02, 0.02) [0.93] | -0.05 (-0.11, 0.01) [0.13] |
| CYP2C19 RM | -0.04 (-0.10, 0.02) [0.23] | -0.03 (-0.07, 0.02) [0.26] | 0.00 (-0.03, 0.02) [0.66] | -0.01 (-0.07, 0.05) [0.78] |
| CYP2C19 UM | -0.06 (-0.17, 0.06) [0.36] | -0.03 (-0.12, 0.06) [0.48] | -0.04 (-0.08, -0.01) [0.025] | 0.02 (-0.11, 0.14) [0.79] |
| **Citalopram/Escitalopram** |  |  |  |  |
| CYP2C19 PM | 0.08 (-0.05, 0.21) [0.25] | 0.05 (-0.05, 0.15) [0.35] | 0.03 (-0.02, 0.08) [0.20] | 0.03 (-0.10, 0.17) [0.65] |
| CYP2C19 IM | -0.02 (-0.07, 0.04) [0.54] | -0.02 (-0.06, 0.02) [0.42] | 0.01 (-0.01, 0.02) [0.54] | -0.01 (-0.07, 0.04) [0.67] |
| CYP2C19 RM | -0.02 (-0.08, 0.04) [0.51] | -0.03 (-0.07, 0.02) [0.23] | 0.02 (-0.00, 0.04) [0.08] | -0.05 (-0.11, 0.01) [0.10] |
| CYP2C19 UM | -0.05 (-0.17, 0.06) [0.38] | -0.04 (-0.13, 0.05) [0.37] | 0.01 (-0.03, 0.05) [0.59] | -0.08 (-0.20, 0.04) [0.20] |
| **Sertraline** |  |  |  |  |
| CYP2C19 PM | 0.09 (-0.19, 0.36) [0.53] | 0.06 (-0.16, 0.27) [0.61] | 0.01 (-0.08, 0.01) [0.88] | 0.01 (-0.27, 0.30) [0.92] |
| CYP2C19 IM | -0.01 (-0.13, 0.11) [0.85] | -0.01 (-0.11, 0.08) [0.76] | 0.05 (0.01, 0.09) [0.007] | -0.17 (-0.29, -0.05) [0.007] |
| CYP2C19 RM | -0.01 (-0.13, 0.12) [0.93] | -0.01 (-0.11, 0.09) [0.88] | 0.03 (-0.02, 0.07) [0.24] | -0.07 (-0.20, 0.06) [0.32] |
| CYP2C19 UM | -0.12 (-0.38, 0.14) [0.37] | -0.07 (-0.28, 0.13) [0.47] | 0.05 (-0.04, 0.13) [0.29] | -0.24 (-0.51, 0.03) [0.08] |

LDL, low-density lipoprotein; HDL, high-density lipoprotein; PM, poor metaboliser; IM, intermediate metaboliser; RM, rapid metaboliser; UM, ultra-rapid metaboliser.

**Figure 3.**
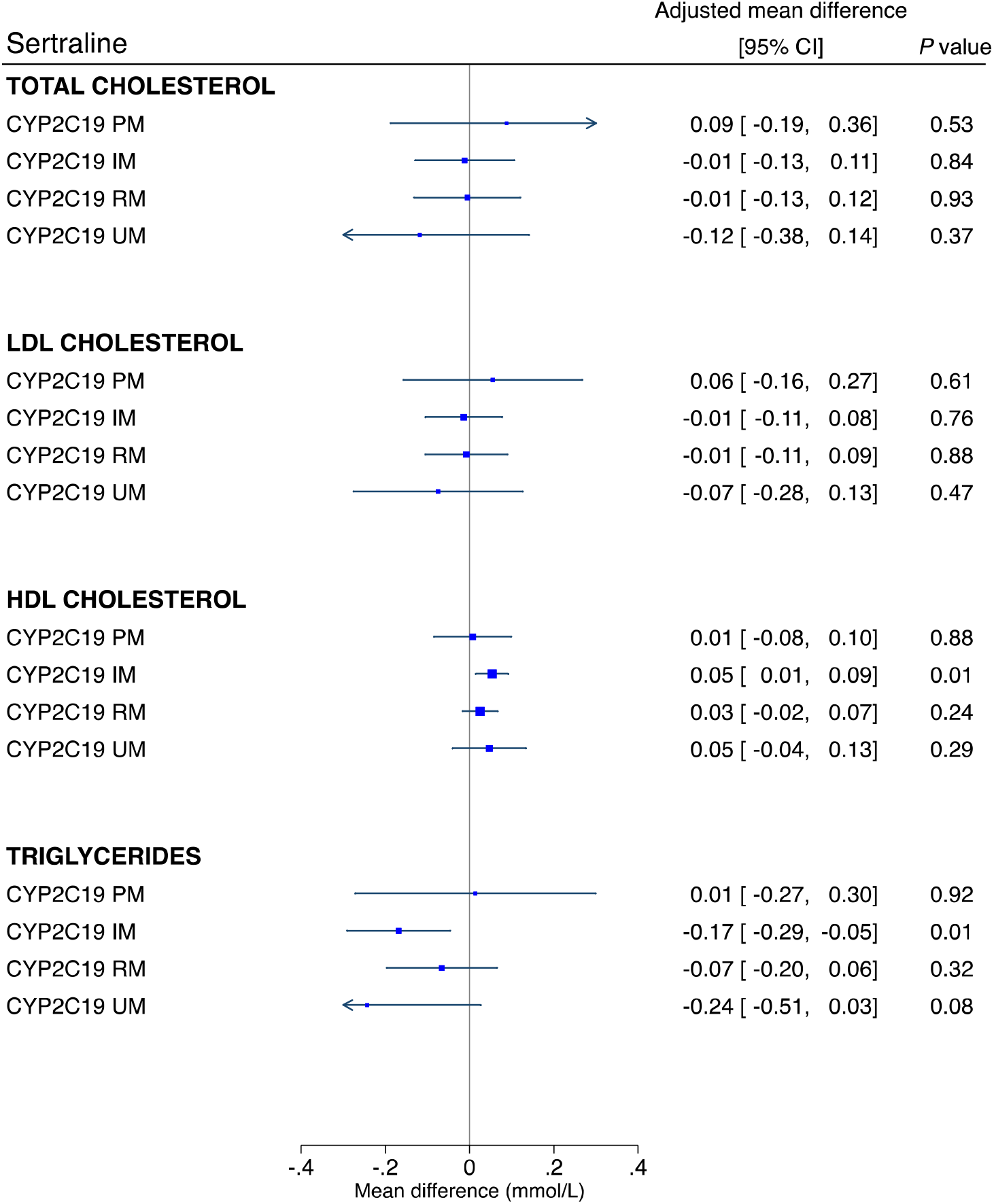
The influence of CYP2C19 metabolic phenotypes on lipid parameters in participants taking sertraline. PM, poor metaboliser; IM intermediate metaboliser; RM, rapid metaboliser; UM, ultra-rapid metaboliser; LDL, low-density lipoprotein; HDL, high-density lipoprotein; mmol/L, millimoles per litre. Linear regression models were adjusted for age (continuous), sex (binary), genetically-determined ancestry group (categorical), concomitant use of cholesterol lowering medications (binary) and use of strong/moderate CYP2C19 inhibitors (binary). The CYP2C19 normal metaboliser phenotype is the reference group.

No significant pharmacogenetic associations were found for other antidepressants or for antipsychotics. Use of strong/moderate *CYP2C19* inhibitors was not a significant predictor of any lipid parameter in CYP2C19 substrates. Use of strong/moderate *CYP2D6* inhibitors was associated with higher triglyceride levels in participants taking antipsychotics (0·29 mmol/L, 95% CI: 0·14 to 0·44, *p*=<0·001, n=277). For venlafaxine, although *CYP2D6* inhibitor use was significant for total cholesterol and LDL-C, this subgroup was too small to meaningfully interpret (n=16).

## DISCUSSION

In this population-based, observational, cohort study using genetic and cross-sectional data on 469,739 participants from UK Biobank, we found that the use of amitriptyline, citalopram/escitalopram, fluoxetine, paroxetine, sertraline, and venlafaxine were each all significantly associated with adverse levels of total cholesterol, LDL-C, HDL-C and triglycerides. In participants taking sertraline, we found that the *CYP2C19* intermediate metaboliser phenotype was significantly associated with higher HDL-C and lower triglyceride levels. Antipsychotic use was significantly associated with lower HDL-C and increased triglycerides.

In contrast with a previous review,^12^ venlafaxine was the antidepressant associated with the worst lipid profile in our study - with the highest levels of total cholesterol, LDL-C, triglycerides, and greatest TC:HDL ratio. A similar profile was observed for paroxetine and sertraline, but, apart from HDL-C, the range of the CIs were generally less favourable for venlafaxine. Amitriptyline was associated with more modest differences in total cholesterol and LDL-C but was associated with the lowest HDL-C levels. Citalopram/escitalopram appeared to have the least detrimental lipid profile - only fluoxetine was associated with a point estimate indicating less of a reduction in HDL-C (but with very similar CIs), though fluoxetine was associated with higher levels of the other lipids. These results are consistent with some aspects of the 2021 Maudsley Prescribing Guidelines in Psychiatry,^37^ but the latter highlight only venlafaxine, sertraline, and mirtazapine (not studied here) as antidepressants raising total cholesterol, and venlafaxine and mirtazapine as raising LDL-C. No antidepressants are noted to impact HDL-C or triglycerides (notable given the largest effects in this study were observed for triglycerides, which contributes to metabolic syndrome).

These results suggest that more antidepressants than traditionally thought are associated with adverse lipid profiles. Considering the magnitude of associations (and their associated CIs) identified in our study, antidepressant choice may be most clinically relevant for patients at risk for cardiovascular morbidity, including those with severe mental illness.^2^ Policymakers and guideline panels should consider whether the introduction of baseline and regular (e.g. annual) monitoring of lipids is warranted in those prescribed antidepressants. A greater amount of research efforts have been put into elucidating the cardiometabolic effects of antipsychotics, where baseline and annual monitoring is already recommended in NICE guidelines.^7^ Given that antidepressants are some of the most commonly prescribed medications - almost 80 million were prescribed to over 7.8 million people in England alone in 2020/2021 (with numbers increasing year-on-year)^38^ - it is paramount to fully understand these effects and their determinants (genetic and environmental) in order to minimise adverse reactions for patients. Antidepressants in our study reflect contemporary prescribing in England,^39^ but further research into the effects of others (e.g. mirtazapine, duloxetine, trazadone) is needed.

Our pharmacogenetic results suggest that, in people taking sertraline, the *CYP2C19* intermediate metaboliser phenotype could be protective for HDL-C, possibly offsetting the overall lower HDL-C associated with sertraline and may also limit the higher triglyceride levels associated with sertraline (noting, however, the lower end of the CIs were close to the null). The reasons for these associations, which require replication, are unclear. A priori, we hypothesized that the presence of one or more low function *CYP2D6* or *CYP2C19* alleles would be associated with increased risk of adverse drug reactions such as altered lipid profile. A similar paradoxical finding, of less adverse events in those with reduced *CYP2C19* metabolic activity taking sertraline, has also been reported in a paediatric sample (though lipids were not reported).^40^ For sertraline, post-hoc analyses identified a statistical interaction of the *CYP2C19* intermediate metaboliser phenotype and use of cholesterol lowering medications, with the effect observed in participants not taking cholesterol lowering medications. Given increasing polypharmacy and high co-prescription rate of antidepressants and cholesterol lowering medications, further research into possible interactions, both drug-drug and drug-drug-gene, on clinical outcomes (e.g., depression, dyslipidaemia) and adverse reactions is warranted. Nevertheless, any impact of *CYP2D6* and *CYP2C19* on lipids in people taking antidepressants/antipsychotics may be relatively small - research into other genes (e.g. *HTR2A*,^11^ *NCAM1* and *KIAA1211*^41^) could be informative, as well as research into other possible biological mechanisms of why antidepressants adversely impact lipids.

We were surprised not to find an association between antipsychotic use and total cholesterol or LDL-C, given that this relationship is well-established.^4–7,13,37^ This could have been due to the population-based sampling used, which resulted in a relatively low number of participants taking each antipsychotic - no individual antipsychotic reached the ≥1,800-participant threshold. Our analyses therefore considered all antipsychotics as one potentially heterogenous group – post-hoc analyses, however, excluding the most frequently reported antipsychotic, prochlorperazine (commonly used for nausea), was consistent. It is also likely that many participants taking antipsychotics in our study were not taking them for psychosis or bipolar disorder, but rather for nausea, anxiety, hiccups, and therefore may have been on substantially lower doses. Another explanation could relate to clinical cardiovascular management. Almost 30% of participants taking antipsychotics also took cholesterol lowering medications, nearly double than in the group not taking antipsychotics/antidepressants. Given known cardiometabolic adverse reactions, and the role of cardiovascular morbidity in the premature mortality in this population, UK general practitioners and psychiatrists have placed particular emphasis on managing cardiovascular risks and promoting health behaviours, including the prescription of statins and promotion of smoking cessation, a healthy diet and exercise. This area has also been the subject of UK government public health initiatives^42^ and interventional research.^43^ It is possible that closer monitoring could be limiting the detrimental effects of antipsychotics (noting participants were recruited 2006-2010); different results may be observed in other settings.

This study has several strengths. We included a very large sample of participants from UK Biobank - enabling robust comparisons, particularly across individual antidepressants. Our pharmacogenetic analyses did not exclude participants of non-European ancestry, a practice common in genetic studies.^44^ Analyses were adjusted to account for key co-variates (e.g. cholesterol lowering medications and *CYP2D6*/*CYP2C19* inhibitors). To our knowledge, this is also the first large study to investigate CYP450 metabolic phenotypes in this context.

This study also has limitations. Data on medication was self-reported and, along with lipid parameters, measured at one time-point. Data on doses prescribed, medication plasma concentrations and duration of therapy would have enabled more sophisticated analyses. Future studies could consider other lipid parameters (e.g., very-low density lipoprotein) and randomised studies, considering other potential environmental confounders (e.g., diet, physical activity) and with multiple sampling time-points, would provide more definitive evidence. It is also possible that certain psychiatric conditions or symptoms impact lipid parameters directly^8,12^ and/or interact with medications. Despite the very large sample size, the number of non-normal metabolisers (particularly poor metabolisers) was much more modest – as expected, given the prevalence of polymorphisms that result in non-normal phenotypes, limiting statistical power and precision in our pharmacogenetic analyses. Future studies will need to employ different methods, such as oversampling, to ensure inclusion of larger numbers of non-normal metabolisers. We were unable to define *CYP2D6* ultra-rapid metabolisers (any such individuals are treated as normal metabolisers by default in our analyses) - however, the prevalence of this phenotype is very rare, and any impact, likely minimal.

Around 94% of our sample were of white ethnicity and, when compared to the 2011 population estimate (86%),^45^ this highlights that all other ethnicities were under-represented, potentially limiting generalizability, especially as cardiovascular risk is greater in some non-white ethnicities. Greater efforts are required to improve diversity in research.

### Conclusion

Antidepressants, one of the most commonly prescribed class of medications,^38^ are significantly associated with adverse lipid profiles - potentially warranting the introduction of baseline and regular monitoring of lipids, in a similar way to what NICE already recommends for antipsychotics.^7^ Venlafaxine was associated with the worst lipid profile whilst citalopram/escitalopram had the smallest effect sizes for raised lipids. Further research should investigate why the *CYP2C19* intermediate metaboliser phenotype may be protective for HDL-C and triglycerides in people taking sertraline.

Antipsychotic use was not associated with total cholesterol or LDL-C in our sample, possibly due to heterogeneity, modest statistical power, and/or co-prescribed cholesterol lowering medication, but was associated with lower HDL-C and increased triglycerides.

## Data Availability

All data used in this study is publicly available to authorised researchers via the UK Biobank.

https://www.ukbiobank.ac.uk/

## STATEMENTS

## Acknowledgements

This research has been conducted using data from UK Biobank, a major biomedical database. We thank all of the UK Biobank participants for giving their time to participate in the research and the UK Biobank for providing access to the data.

## Funding

ARB is funded by the Wellcome Trust through a PhD Fellowship in Mental Health Science. This research was funded in whole or in part by the Wellcome Trust. For the purpose of Open Access, the author has applied a CC BY public copyright licence to any Author Accepted Manuscript (AAM) version arising from this submission.

EB acknowledges the support of: Medical Research Council (G1100583, MR/W020238/1), National Institute of Health Research (NIHR200756), Mental Health Research UK - John Grace QC Scholarship 2018, Economic Social Research Council’s Co-funded doctoral award, The British Medical Association’s Margaret Temple Fellowship, Medical Research Council New Investigator and Centenary Awards (G0901310, G1100583), NIHR Biomedical Research Centre at University College London Hospitals NHS Foundation Trust and University College London.

BW is funded by the China Scholarship Council-UCL Joint Research Scholarship.

DPJO is supported by the University College London Hospitals NIHR Biomedical Research Centre and the NIHR North Thames Applied Research Collaboration. This funder had no role in study design, data collection, data analysis, data interpretation, or writing of the report. The views expressed in this article are those of the authors and not necessarily those of the NHS, the NIHR, or the Department of Health and Social Care.

## Declaration of Interest

There are no conflicts of interest.

## Author Contributions

EB, ARB, IAZ, BW conceived the study. ARB, IAZ, EB analysed the data. ARB, IAZ, BW, EZ, MC, CG, NSK, YW, MW, YD, DO, EB interpreted the data. ARB drafted the manuscript, supervised by EB, and all authors critically reviewed the manuscript for important intellectual content and approved the final version. ARB, IAZ, BW, EZ, EB had full access to the data.

## Data Availability

All data used in this study is publicly available to authorised researchers via the UK Biobank. This study was conducted under UK Biobank application ID 20737. Detail on the available data can be found here: https://biobank.ndph.ox.ac.uk/showcase.

## Notes

### Competing Interest Statement

The authors have declared no competing interest.

### Author Declarations

UK Biobank received ethical approval from the North West - Haydock Research Ethics Committee (reference: 21/NW/0157).

