## Supplementary Material for "Associations of antidepressants and antipsychotics with lipid parameters: Do *CYP2D6*/*CYP2C19* genes play a role? A UK population-based study"

Supplementary Table 1. Extended sample characteristics.

|  | **No antidepressant/ antipsychotic** | **Amitriptyline** | **Fluoxetine** | **Citalopram/ Escitalopram** | **Sertraline** | **Paroxetine** | **Venlafaxine** | **Any Antipsychotic** |
| --- | --- | --- | --- | --- | --- | --- | --- | --- |
| **N** | 431,853 | 9,423 | 6,223 | 9,837 | 2,324 | 2,152 | 2,131 | 3,255 |
| **Age (years), median (IQR)** | 58 (50, 63) | 60 (53, 64) | 55 (48, 61) | 56 (49, 61) | 56 (49, 62) | 58 (52, 63) | 56 (49, 62) | 57 (50, 63) |
| **Sex, n (%)** |  |  |  |  |  |  |  |  |
| Male | 202,942 (47.0) | 2,770 (29.4) | 1,724 (27.7) | 3,037 (30.9) | 773 (33.3) | 742 (34.5) | 755 (35.4) | 1,369 (42.1) |
| Female | 228,911 (53.0) | 6,653 (70.6) | 4,499 (72.3) | 6,800 (69.1) | 1,551 (66.7) | 1,410 (65.5) | 1,376 (64.6) | 1,886 (57.9) |
| **Ethnic background (self-reported), n (%)** |  |  |  |  |  |  |  |  |
| Asian | 8,647 (2.0) | 130 (1.4) | 46 (0.7) | 135 (1.4) | 24 (1.0) | 21 (1.0) | 14 (0.7) | 74 (2.3) |
| Black | 6,966 (1.6) | 102 (1.1) | 46 (0.7) | 61 (0.6) | 11 (0.5) | 14 (0.7) | 15 (0.7) | 83 (2.6) |
| Mixed | 2,490 (0.6) | 60 (0.6) | 41 (0.7) | 71 (0.7) | 20 (0.9) | 7 (0.3) | 8 (0.4) | 38 (1.2) |
| White | 406,403 (94.1) | 9,008 (95.6) | 6,024 (96.8) | 9,454 (96.1) | 2,245 (96.6) | 2,090 (97.1) | 2,056 (96.5) | 2,993 (92.0) |
| Other | 5,308 (1.2) | 84 (0.9) | 44 (0.7) | 73 (7) | 19 (0.8) | 7 (0.3) | 21 (1.0) | 9 (1.5) |
| Not stated | 2,039 (0.5) | 39 (0.4) | 22 (0.4) | 43 (0.4) | 5 (0.2) | 13 (0.6) | 17 (0.8) | 18 (0.5) |
| **BMI (kg/m2), median (IQR)** | 26.7 (24.1, 29.7) | 28.0 (25.0, 31.9) | 27.9 (24.9, 31.8) | 27.7 (24.6, 31.6) | 27.8 (24.8, 31.5) | 28.1 (25.1, 31.8) | 28.4 (25.4, 32.0) | 28.2 (25.1, 32.1) |
| **BMI (kg/m2) categories, n (%)** |  |  |  |  |  |  |  |  |
| <18.5 (underweight) | 2,188 (0.5) | 44 (0.5) | 29 (0.5) | 58 (0.6) | 16 (0.7) | 10 (0.5) | 10 (0.5) | 26 (0.8) |
| 18.5-24.9 (healthy weight) | 142,649 (33.0) | 2,303 (24.4) | 1,568 (25.2) | 2,685 (27.3) | 591 (25.4) | 506 (23.5) | 452 (21.2) | 766 (23.5) |
| 25-29.9 (overweight) | 184,394 (42.7) | 3,651 (38.8) | 2,437 (39.2) | 3,719 (37.8) | 915 (39.4) | 855 (39.7) | 840 (39.4) | 1,227 (37.7) |
| >30 (obese) | 101,005 (23.4) | 3,327 (35.3) | 2,152 (34.6) | 3,333 (33.9) | 781 (33.6) | 767 (35.6) | 818 (38.4) | 1,208 (37.1) |
| Missing | 1,617 (0.4) | 98 (1.0) | 37 (0.6) | 42 (0.4) | 21 (0.9) | 14 (0.7) | 11 (0.5) | 28 (0.9) |
| **Self-reported illnesses, n (%)** |  |  |  |  |  |  |  |  |
| Depression | 7,625 (1.8) | 1,651 (17.5) | 4,378 (70.4) | 6,480 (65.9) | 1,419 (61.1) | 1,327 (61.7) | 1,565 (73.4) | 1,031 (31.7) |
| Anxiety | 2,434 (0.6) | 434 (4.6) | 594 (9.5) | 1,457 (14.8) | 221 (9.5) | 446 (20.7) | 213 (10.0) | 274 (8.4) |
| Schizophrenia | 74 (<0.1) | 9 (0.1) | 22 (1.0) | 40 (0.4) | 25 (1.1) | 7 (0.3) | 27 (1.3) | 557 (17.1) |
| Bipolar disorder/mania/manic depression | 483 (0.1) | 43 (0.5) | 83 (1.3) | 135 (1.4) | 47 (2.0) | 50 (2.3) | 89 (4.2) | 466 (14.3) |
| **Cholesterol lowering medication use, n (%)** | 72,371 (16.8) | 2,704 (28.7) | 1,333 (21.4) | 2,137 (21.7) | 564 (24.3) | 580 (27.0) | 507 (23.8) | 885 (27.2) |
| **Total cholesterol (mmol/L), mean (SD)** | 5.69 (1.14) | 5.68 (1.26) | 5.81 (1.21) | 5.70 (1.17) | 5.78 (1.23) | 5.79 (1.22) | 5.83 (1.22) | 5.55 (1.25) |
| **Total cholesterol (mmol/L) categories, n (%)** |  |  |  |  |  |  |  |  |
| ≤5 | 119,539 (27.7) | 2,874 (30.5) | 1,633 (26.3) | 2,746 (27.9) | 642 (27.6) | 570 (26.5) | 520 (24.4) | 1,104 (33.9) |
| >5 | 312,180 (72.3) | 6,547 (69.5) | 4,588 (73.7) | 7,090 (72.1) | 1,681 (72.3) | 1,580 (73.4) | 1,611 (75.6) | 2,151 (66.1) |
| Missing | 137 (<0.1) | 2 (<0.1) | 2 (<0.1) | 1 (<0.1) | 1 (<0.1) | 2 (0.1) | 0 (0) | 0 (0) |
| **LDL cholesterol (mmol/L), mean (SD)** | 3.56 (0.87) | 3.54 (0.96) | 3.63 (0.92) | 3.56 (0.89) | 3.63 (0.93) | 3.62 (0.92) | 3.66 (0.94) | 3.47 (0.94) |
| **LDL cholesterol (mmol/L), categories, n (%)** |  |  |  |  |  |  |  |  |
| ≤3 | 116,585 (27.0) | 2,880 (30.6) | 1,643 (26.4) | 2,761 (28.1) | 618 (26.6) | 573 (26.6) | 538 (25.3) | 1,061 (32.6) |
| >3 | 314,330 (72.8) | 6,525 (69.3) | 4,562 (73.3) | 7,051 (71.7) | 1,697 (73.0) | 1,574 (73.1) | 1,589 (74.6) | 2,185 (67.1) |
| Missing | 938 (0.2) | 18 (0.2) | 18 (0.3) | 25 (0.3) | 9 (0.4) | 5 (0.2) | 4 (0.2) | 9 (0.3) |
| **HDL cholesterol (mmol/L), mean (SD)** | 1.45 (0.38) | 1.41 (0.38) | 1.45 (0.39) | 1.44 (0.38) | 1.41 (0.38) | 1.42 (0.37) | 1.41 (0.39) | 1.35 (0.37) |
| **HDL cholesterol (mmol/L), categories, n (%)** |  |  |  |  |  |  |  |  |
| ≥1 | 357,662 (82.8) | 7,475 (79.3) | 5,109 (82.1) | 7,963 (81.0) | 1,843 (79.3) | 1,724 (80.1) | 1,700 (79.8) | 2,464 (75.7) |
| <1 | 37,706 (8.7) | 1,089 (11.6) | 577 (9.3) | 984 (10.0) | 272 (11.7) | 219 (10.2) | 237 (11.1) | 481 (14.8) |
| Missing | 36,485 (8.5) | 859 (9.1) | 537 (8.6) | 890 (9.1) | 209 (9.0) | 209 (9.7) | 194 (9.1) | 310 (9.5) |
| **Triglycerides (mmol/L), mean (SD)** | 1.73 (1.01) | 2.00 (1.15) | 1.91 (1.14) | 1.88 (1.13) | 1.98 (1.15) | 2.06 (1.23) | 2.07 (1.24) | 2.05 (1.26) |
| **Triglycerides (mmol/L), categories, n (%)** |  |  |  |  |  |  |  |  |
| ≤2.3 | 341,837 (79.2) | 6,652 (70.6) | 4,565 (73.4) | 7,364 (74.9) | 1,648 (70.9) | 1,471 (68.4) | 1,462 (68.6) | 2,256 (69.3) |
| >2.3 | 89,533 (20.7) | 2,759 (29.3) | 1,655 (26.6) | 2,460 (25.0) | 673 (29.0) | 681 (31.6) | 665 (31.2) | 994 (30.5) |
| Missing | 483 (0.1) | 12 (0.1) | 3 (0.1) | 13 (0.1) | 3 (0.1) | 0 (0) | 4 (0.2) | 5 (0.2) |
| **TC:HDL ratio, mean (SD)** | 4.12 (1.12) | 4.23 (1.20) | 4.20 (1.18) | 4.18 (1.17) | 4.32 (1.23) | 4.28 (1.19) | 4.37 (1.24) | 4.33 (1.27) |
| **N with genetic data passing quality control** | ·· | 8,308 | 5,510 | 8,705 | 1,981 | 1,935 | 1,906 | 2,781 |
| **Genetic ancestry group, n (%)** |  |  |  |  |  |  |  |  |
| Mixed | ·· | 119 (1.4) | 80 (1.5) | 141 (1.6) | 26 (1.3) | 23 (1.2) | 21 (1.1) | 62 (2.2) |
| Mixed-European | ·· | 173 (2.1) | 136 (2.5) | 229 (2.6) | 69 (3.5) | 59 (3.0) | 54 (2.8) | 72 (2.6) |
| African | ·· | 129 (1.6) | 56 (1.0) | 84 (1.0) | 16 (0.8) | 13 (0.7) | 17 (0.9) | 111 (4.0) |
| East Asian | ·· | 7 (0.1) | 9 (0.2) | 11 (0.1) | 4 (0.2) | 1 (0.1) | 1 (0.1) | 11 (0.4) |
| European | ·· | 7,754 (93.3) | 5,190 (94.2) | 8,121 (93.3) | 1,842 (93.0) | 1,822 (94.2) | 1,797 (94.3) | 2,457 (88.4) |
| South Asian | ·· | 126 (1.5) | 39 (0.7) | 119 (1.4) | 24 (1.2) | 17 (0.9) | 16 (0.8) | 68 (2.5) |
| **CYP2D6 metabolic phenotype,^1^ n (%)** |  |  |  |  |  |  |  |  |
| Normal metaboliser | ·· | 5,916 (71.2) | 3,928 (71.3) | 6,228 (71.6) | ·· | 1,368 (70.7) | 1,368 (71.8) | 1,958 (70.4) |
| Intermediate metaboliser | ·· | 1,961 (23.6) | 1,278 (23.2) | 2,007 (23.1) | ·· | 457 (23.6) | 433 (22.7) | 682 (24.5) |
| Poor metaboliser | ·· | 431 (5.2) | 304 (5.5) | 470 (5.4) | ·· | 110 (5.7) | 105 (5.5) | 141 (5.1) |
| **Strong/moderate CYP2D6 inhibitor(s) use, n (%)** | ·· | 342 (4.1) | 5,510 (100)^2^ | 89 (1.0) | ·· | 1,935 (100)^2^ | 16 (0.8) | 277 (10.0) |
| **CYP2C19 metabolic phenotype,^1^ n (%)** |  |  |  |  |  |  |  |  |
| Ultra-rapid metaboliser | ·· | 366 (4.4) | ·· | 361 (4.2) | 76 (3.8) | ·· | ·· | ·· |
| Rapid metaboliser | ·· | 1,993 (24.0) | ·· | 2,180 (25.0) | 476 (24.0) | ·· | ·· | ·· |
| Normal metaboliser | ·· | 3,162 (38.1) | ·· | 3,338 (38.4) | 767 (38.7) | ·· | ·· | ·· |
| Intermediate metaboliser | ·· | 2,519 (30.3) | ·· | 2,555 (29.4) | 595 (30.0) | ·· | ·· | ·· |
| Poor metaboliser | ·· | 268 (3.2) | ·· | 271 (3.11) | 67 (3.4) | ·· | ·· | ·· |
| **Strong/moderate CYP2C19 inhibitor(s) use, n (%)** | ·· | 246 (3.0) | ·· | 55 (0.6) | 23 (1.2) | ·· | ·· | ·· |

BMI, body mass index; mmol/L, millimoles per litre; LDL-C, low-density lipoprotein cholesterol; HDL-C, high-density lipoprotein cholesterol; TC:HDL, total cholesterol to high-density lipoprotein cholesterol.

^1^ Medications were defined as CYP2D6 and/or CYP2C19 substrates in accordance with the Clinical Pharmacogenetics Implementation Consortium (CPIC).^14,15,34^

^2^ Fluoxetine and paroxetine are defined as strong inhibitors for CYP2D6.^33^

Supplementary Table 2. Unadjusted lipid parameters stratified by concomitant cholesterol lowering medication status.

|  | **No antidepressant/ antipsychotic** | **Amitriptyline** | **Fluoxetine** | **Citalopram/ Escitalopram** | **Sertraline** | **Paroxetine** | **Venlafaxine** | **Any Antipsychotic** |
| --- | --- | --- | --- | --- | --- | --- | --- | --- |
| **N** | 431,853 | 9,423 | 6,223 | 9,837 | 2,324 | 2,152 | 2,131 | 3,255 |
| **n (%) taking cholesterol lowering medication** | 72,371 (16.8) | 2,704 (28.7) | 1,333 (21.4) | 2,137 (21.7) | 564 (24.3) | 580 (27.0) | 507 (23.8) | 885 (27.2) |
| **Total cholesterol (mmol/L), mean (SD)** |  |  |  |  |  |  |  |  |
| *Taking cholesterol lowering medications* | 4.73 (1.00) | 4.75 (1.04) | 4.91 (1.08) | 4.84 (1.05) | 4.93 (1.03) | 4.98 (1.13) | 4.97 (1.13) | 4.70 (1.06) |
| *No cholesterol lowering medications* | 5.88 (1.06) | 6.06 (1.14) | 6.05 (1.12) | 5.94 (1.09) | 6.05 (1.16) | 6.09 (1.12) | 6.10 (1.12) | 5.87 (1.16) |
| **LDL cholesterol (mmol/L), mean (SD)** |  |  |  |  |  |  |  |  |
| *Taking cholesterol lowering medications* | 2.82 (0.73) | 2.83 (0.75) | 2.95 (0.78) | 2.90 (0.76) | 2.97 (0.74) | 2.99 (0.83) | 2.98 (0.82) | 2.80 (0.75) |
| *No cholesterol lowering medications* | 3.70 (0.81) | 3.83 (0.87) | 3.81 (0.87) | 3.74 (0.84) | 3.84 (0.89) | 3.85 (0.85) | 3.87 (0.87) | 3.72 (0.87) |
| **HDL cholesterol (mmol/L), mean (SD)** |  |  |  |  |  |  |  |  |
| *Taking cholesterol lowering medications* | 1.31 (0.35) | 1.28 (0.35) | 1.30 (0.35) | 1.29 (0.35) | 1.28 (0.34) | 1.31 (0.35) | 1.28 (0.34) | 1.24 (0.34) |
| *No cholesterol lowering medications* | 1.48 (0.38) | 1.46 (0.38) | 1.50 (0.39) | 1.47 (0.38) | 1.45 (0.39) | 1.46 (0.37) | 1.45 (0.39) | 1.39 (0.38) |
| **Triglycerides (mmol/L), mean (SD)** |  |  |  |  |  |  |  |  |
| *Taking cholesterol lowering medications* | 1.93 (1.09) | 2.16 (1.20) | 2.19 (1.27) | 2.14 (1.21) | 2.21 (1.23) | 2.28 (1.30) | 2.43 (1.43) | 2.28 (1.37) |
| *No cholesterol lowering medications* | 1.69 (0.99) | 1.93 (1.13) | 1.83 (1.08) | 1.80 (1.10) | 1.90 (1.12) | 1.97 (1.20) | 1.96 (1.16) | 1.97 (1.20) |

LDL, low-density lipoprotein; HDL, high-density lipoprotein.

Supplementary Table 3. Adjusted associations of antidepressants and antipsychotics with lipid parameters.

|  | **Amitriptyline** | **Fluoxetine** | **Citalopram/ Escitalopram** | **Sertraline** | **Paroxetine** | **Venlafaxine** | **Any Antipsychotic** |
| --- | --- | --- | --- | --- | --- | --- | --- |
| **N taking the medication** | 9,423 | 6,223 | 9,837 | 2,324 | 2,152 | 2,131 | 3,255 |
| **Total cholesterol (mmol/L)** |  |  |  |  |  |  |  |
| Effect estimate (95% CI) | 0.05  (0.03, 0.07) | 0.16 (0.13, 0.19) | 0.06  (0.04, 0.08) | 0.16 (0.12, 0.21) | 0.17 (0.12, 0.21) | 0.21 (0.17, 0.26) | -0.03 (-0.06, 0.01) |
| **LDL cholesterol (mmol/L)** |  |  |  |  |  |  |  |
| Effect estimate (95% CI) | 0.06 (0.04, 0.08) | 0.13 (0.11, 0.15) | 0.06  (0.04, 0.07) | 0.15 (0.11, 0.18) | 0.14  (0.10, 0.17) | 0.17 (0.14, 0.21) | 0.01 (-0.02, 0.03) |
| **HDL cholesterol (mmol/L)** |  |  |  |  |  |  |  |
| Effect estimate (95% CI) | -0.08 (-0.09, -0.08) | -0.04  (-0.05, -0.03) | -0.05 (-0.05, -0.04) | -0.06  (-0.08, -0.05) | -0.05  (-0.07, -0.04) | -0.06  (-0.07, -0.04) | -0.10 (-0.11, -0.08) |
| **Triglycerides (mmol/L)** |  |  |  |  |  |  |  |
| Effect estimate (95% CI) | 0.30 (0.28, 0.32) | 0.24 (0.22, 0.27) | 0.20 (0.18, 0.22) | 0.28 (0.24, 0.32) | 0.34 (0.30, 0.38) | 0.36 (0.32, 0.41) | 0.31 (0.28, 0.35) |
| **TC:HDL ratio** |  |  |  |  |  |  |  |
| Effect estimate (95% CI) | 0.28  (0.26, 0.30) | 0.23  (0.20, 0.25) | 0.18  (0.16, 0.21) | 0.32  (0.28, 0.37) | 0.28  (0.24, 0.33) | 0.35  (0.31, 0.40) | 0.28  (0.25, 0.32) |

LDL, low-density lipoprotein; HDL, high-density lipoprotein.

All linear regression models were adjusted for age, sex and concomitant use of cholesterol lowering medications; effect estimates are coefficients for the main predictor variable, which was a binary variable defined by whether participants were taking the relevant medication (or not). A total of 469,591 participants contributed total cholesterol data, 468,708 for LDL cholesterol, 429,873 for HDL cholesterol and 469,216 for triglycerides. All coefficients were significant at the p<0.001 level, except for antipsychotics x total cholesterol (p=0.137) and antipsychotics x LDL cholesterol (p=0.598).

Supplementary Table 4. The influence of *CYP2D6* metabolic phenotypes on lipid parameters in participants taking antidepressants or antipsychotics.

|  | **Effect estimate (95% CI) [*p* value]** | | | |
| --- | --- | --- | --- | --- |
| ***Predictors*** | **Total cholesterol (mmol/L)** | **LDL cholesterol (mmol/L)** | **HDL cholesterol (mmol/L)** | **Triglycerides (mmol/L)** |
| **Amitriptyline** |  |  |  |  |
| *CYP2D6 PM* | 0.01 (-0.10, 0.12) [0.83] | 0.01 (-0.07, 0.09) [0.81] | 0.00 (-0.03, 0.04) [0.94] | 0.04 (-0.07, 0.16) [0.44] |
| *CYP2D6 IM* | 0.02 (-0.04, 0.07) [0.54] | 0.01 (-0.03, 0.06) [0.55] | -0.01 (-0.03, 0.01) [0.49] | 0.02 (-0.04, 0.08) [0.48] |
| ***Fluoxetine*** |  |  |  |  |
| *CYP2D6 PM* | -0.11 (-0.24, 0.02) [0.09] | -0.10 (-0.20, 0.00) [0.05] | 0.022 (-0.022, 0.066) [0.321] | -0.12 (-0.25, 0.01) [0.07] |
| *CYP2D6 IM* | 0.01 (-0.06, 0.08) [0.72] | 0.02 (-0.04, 0.07) [0.56] | -0.004 (-0.027, 0.020) [0.748] | -0.01 (-0.08, 0.06) [0.70] |
| **Citalopram/Escitalopram** |  |  |  |  |
| *CYP2D6 PM* | -0.08 (-0.17, 0.02) [0.14] | -0.03 (-0.11, 0.04) [0.40] | -0.03 (-0.07, 0.00) [0.06] | 0.02 (-0.08, 0.12) [0.73] |
| *CYP2D6 IM* | -0.06 (-0.11, -0.01) [0.027] | -0.05 (-0.09, -0.01) [0.016] | -0.00 (-0.02, 0.02) [0.85] | 0.01 (-0.05, 0.06) [0.78] |
| **Paroxetine** |  |  |  |  |
| *CYP2D6 PM* | 0.14 (-0.07, 0.36) [0.19] | 0.12 (-0.04, 0.29) [0.14] | -0.01 (-0.08, 0.06) [0.68] | 0.11 (-0.13, 0.34) [0.38] |
| *CYP2D6 IM* | -0.05 (-0.17, 0.07) [0.41] | -0.03 (-0.12, 0.05) [0.45] | -0.01 (-0.05, 0.03) [0.72] | -0.02 (-0.15, 0.11) [0.77] |
| **Venlafaxine** |  |  |  |  |
| *CYP2D6 PM* | -0.09 (-0.31, 0.14) [0.45] | -0.10 (-0.27, 0.07) [0.24] | 0.06 (-0.02, 0.14) [0.12] | -0.05 (-0.29, 0.19) [0.68] |
| *CYP2D6 IM* | 0.03 (-0.09, 0.15) [0.62] | 0.02 (-0.07, 0.11) [0.69] | 0.01 (-0.03, 0.05) [0.57] | -0.03(-0.16, 0.10) [0.64] |
| **Antipsychotics** |  |  |  |  |
| *CYP2D6 PM* | -0.15 (-0.34, 0.04) [0.13] | -0.08 (-0.23, 0.06) [0.25] | -0.01 (-0.07, 0.05) [0.80] | -0.17 (-0.38, 0.04) [0.12] |
| *CYP2D6 IM* | 0.05 (-0.04, 0.15) [0.27] | 0.06 (-0.01, 0.13) [0.10] | -0.00 (-0.03, 0.03) [0.89] | 0.01 (-0.10, 0.12) [0.83] |

LDL, low-density lipoprotein; HDL, high-density lipoprotein; PM, poor metaboliser; IM, intermediate metaboliser.

Linear regression models were adjusted for age (continuous), sex (binary), genetically-determined ancestry group (categorical), concomitant use of cholesterol lowering medications (binary) and concomitant use of strong/moderate CYP2D6 or CYP2C19 inhibitors (binary). As fluoxetine and paroxetine are themselves considered strong inhibitors for CYP2D6, the ‘strong/moderate CYP2D6 inhibitor’ variable was not included in those models.

Supplementary Table 5. Post-hoc analyses of the influence of the *CYP2C19* intermediate metaboliser genetic metabolic phenotype on HDL cholesterol and triglycerides in participants taking sertraline.

|  | Effect estimate  (95% confidence interval) [*p* value] | |
| --- | --- | --- |
|  | **HDL cholesterol** | **Triglycerides** |
| **Primary model** |  |  |
| CYP2C19 IM – main effect | 0.05 (0.01, 0.09) [0.01] | -0.17 (-0.29, -0.05) [0.007] |
| **Primary model + interaction term of CYP2C19 metabolic phenotype by cholesterol lowering medication status** | | |
| CYP2C19 IM – main effect | 0.08 (0.03, 0.12) [0.001] | -0.15 (-0.29, -0.01) [0.04] |
| CYP2C19 IM x cholesterol lowering medications – interaction term | -0.10 (-0.19, 0.01) [0.03] | -0.07 (-0.36, 0.21) [0.61] |
| **Primary model, stratified by cholesterol lowering medications** |  |  |
| CYP2C19 IM main effect – participants taking cholesterol lowering medications | -0.01 (-0.08, 0.06) [0.73]  n=126 | -0.22 (-0.48, 0.05) [0.10]  n=137 |
| CYP2C19 IM main effect – participants not taking cholesterol lowering medications | 0.08 (0.03, 0.12) [0.001] n=424 | -0.15 (-0.29, -0.02) [0.03]  n=456 |

HDL, high-density lipoprotein; IM, intermediate metaboliser.
